# Regaining perspective on SARS-CoV-2 molecular tracing and its implications

**DOI:** 10.1101/2020.03.16.20034470

**Authors:** Carla Mavian, Simone Marini, Costanza Manes, Ilaria Capua, Mattia Prosperi, Marco Salemi

## Abstract

During the past three months, a new coronavirus (SARS-CoV-2) epidemic has been growing exponentially, affecting over 100 thousand people worldwide, and causing enormous distress to economies and societies of affected countries. A plethora of analyses based on viral sequences has already been published, in scientific journals as well as through non-peer reviewed channels, to investigate SARS-CoV-2 genetic heterogeneity and spatiotemporal dissemination. We examined all full genome sequences currently available to assess the presence of sufficient information for reliable phylogenetic and phylogeographic studies. Our analysis clearly shows severe limitations in the present data, in light of which any finding should be considered, at the very best, preliminary and hypothesis-generating. Hence the need for avoiding stigmatization based on partial information, and for continuing concerted efforts to increase number and quality of the sequences required for robust tracing of the epidemic.

## Main

In December 2019, a novel coronavirus, SARS-CoV-2, was identified in Wuhan, China, as the etiologic agent of coronavirus disease 2019 (COVID-19), which by March 2020 has already spread across more than 80 countries ^1^. Common symptoms of infection include fever, cough, and shortness of breath, while severe cases are characterized by advanced respiratory distress and pneumonia, often resulting in death ^2^. Soon after the first epidemiological data together with SARS-CoV-2 genetic sequences were made available, a glut of phylogeny-based analyses began to circulate discussing, in scientific papers as well as (social) media, countries that might have been fueling the spread. The implications of misunderstanding the real dynamic of the COVID-19 pandemic are extremely dangerous. Ethnic or social discrimination resulting from unsupported assumptions on viral contagion – often amplified by irresponsible, uncontrollable communications – can be highly damaging for people and countries.

Despite (social) media are often vehicle for fake news and boast news hype, it is also worth noting the tremendous effort of the scientific community to provide free, up-to-date information on ongoing studies, as well as critical evaluations. In particular, the US-based NextStrain ^3^ team has been posting real-time updates on the epidemic tracing by molecular analyses. Several discussions and evidence-based debates on controversial hypotheses on the epidemic have ensued — such as the number of untraced infections in the US, the putative virus introduction in Italy through Germany, and the alleged lineage diversification in China^4^ later criticized ^5^— aided by a concerted and productive effort of internationally-renowned research groups in Edinburgh, Glasgow, and Oxford, UK, among others. Recently, an editorial published on Science ^6^ has also highlighted how unsupported or misleading claims circulating in forums, social media, and even peer-reviewed articles, have been led by a substantial over interpretation of the available data. Hence, the urgency to reframe the current debate in more rigorous scientific terms, and quantitatively evaluate whether sufficient information for reliable phylogenetic and phylogeographic studies currently exists, or which gaps need to be addressed.

## Inhomogeneous sampling and phylogeographic uncertainly

Before carrying out any phylogeny-based analysis of virus evolution and spatiotemporal spread, it is crucial to test the quality of sequence data, since uneven sampling, presence of phylogenetic noise, and absence of temporal signal can affect reliability of the results (e.g. ancestral state reconstructions, molecular clock calibrations) ^7^. We downloaded all SARS-CoV-2 full genome sequences deposited on GISAID (https://www.gisaid.org/) ^8^ to compare the number of high quality genomes sampled per country with the number of confirmed cases at the time of sampling, as well as the country’s total population (Figure 1). As March 10^th^ 2020, 331 genome sequences from 29 countries were available, almost twice as much than just one week earlier (March 3^rd^ 2020, 169 genome sequences from 22 countries). Yet, the sampling per country was inhomogeneous, with confirmed cases clearly uncorrelated to the number of genomes (Spearman correlation = 0.03). Moreover, correlation could only be investigated with confirmed cases (as proxy), since not all affected countries have made publicly available the total number of coronavirus testing performed. Even within the same country, sequenced genomes were usually sampled from few hotspots, not necessarily representative of the whole epidemic in that country. Italy, for example, the second country per confirmed cases, has uploaded so far only two genomes from Lombardy and three from Lazio (Table S1), i.e. only two of the twenty Italian regions, all with confirmed cases^9^. China and USA contributed with 123 and 63 genomes respectively (56% combined), while 13 countries contribute with only one or two samples (6.3% combined). Notably, three countries with less than 400 confirmed cases have provided a substantial number of genomes (Netherlands 28, Australia 18, Singapore 11; 17% combined) (Figure 1).

**Figure 1.**
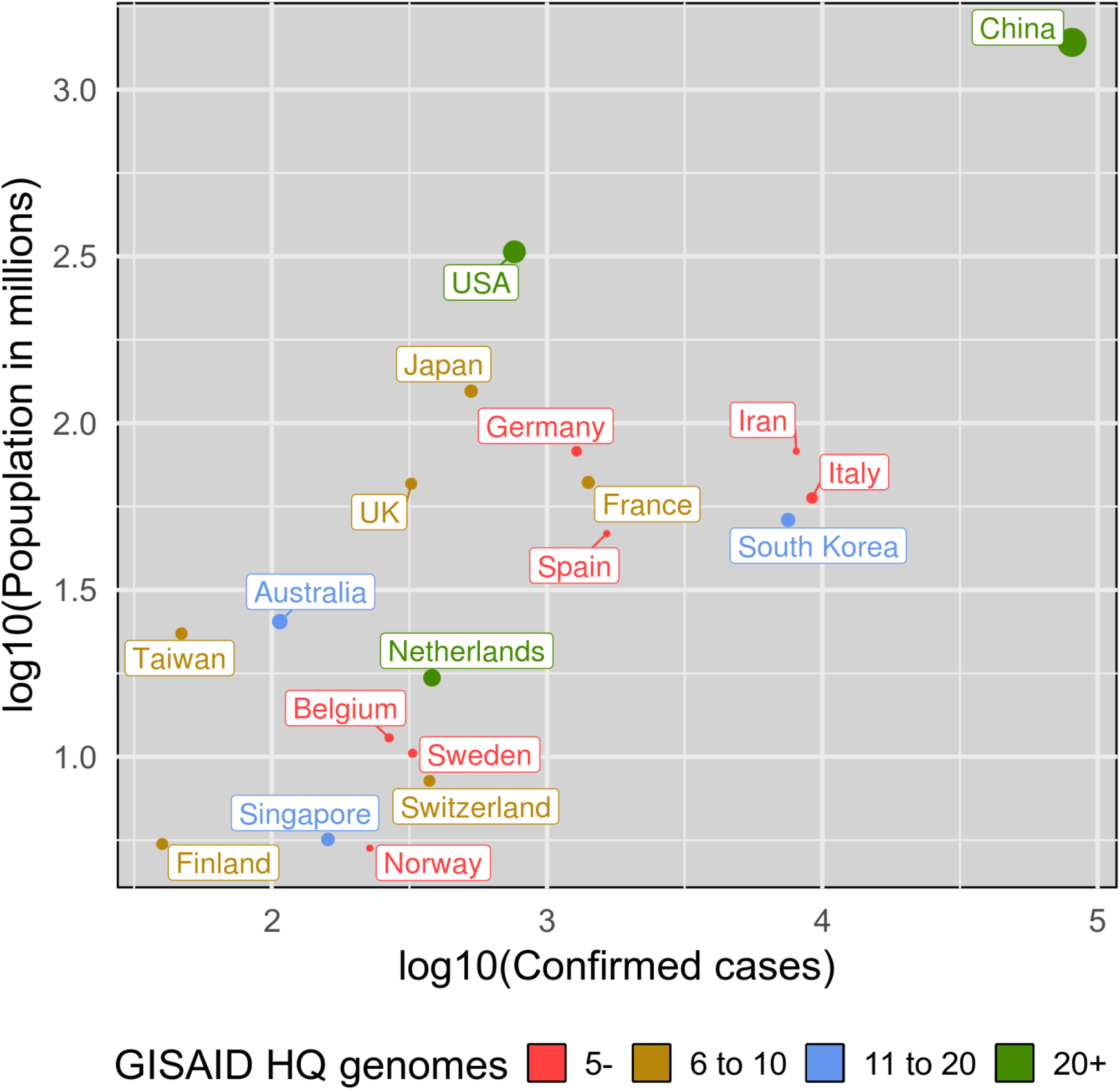
Available SARS-CoV-2 genome sequences and reported cases per country. Selected countries for number of provided high quality GISAID genomes (5 or more), or confirmed cases (200 or more) as of March 10^th^ 2020. On the x axis we have number the confirmed cases (log10 scale), on the y axis the population in millions (log10 scale). The number of genomes determines dot size and color.

The effect of inhomogeneous sampling and missing data on phylogeography reconstructions, like the ones recently rushed through news and (social) media to claim specific dissemination routes of SARS-CoV-2 among countries, can be quite dramatic. An instructive example is the putative introduction of SARS-CoV-2 in Italy from Germany. The maximum likelihood (ML) tree, inferred from the full genome viral sequences available on March 3^rd^, 2020 (Supplementary Figure S1), showed a well-supported cluster of European and Asian sequences (reported in Figure 2a), which contained a subclade (subclade A, Figure 2a) including a sequence isolated in Germany that appears to be paraphyletic (with strong bootstrap support) to an Italian sequence clustering, in turn, with sequences from Finland, Mexico, Germany and Switzerland (Figure 2a).

**Figure 2.**
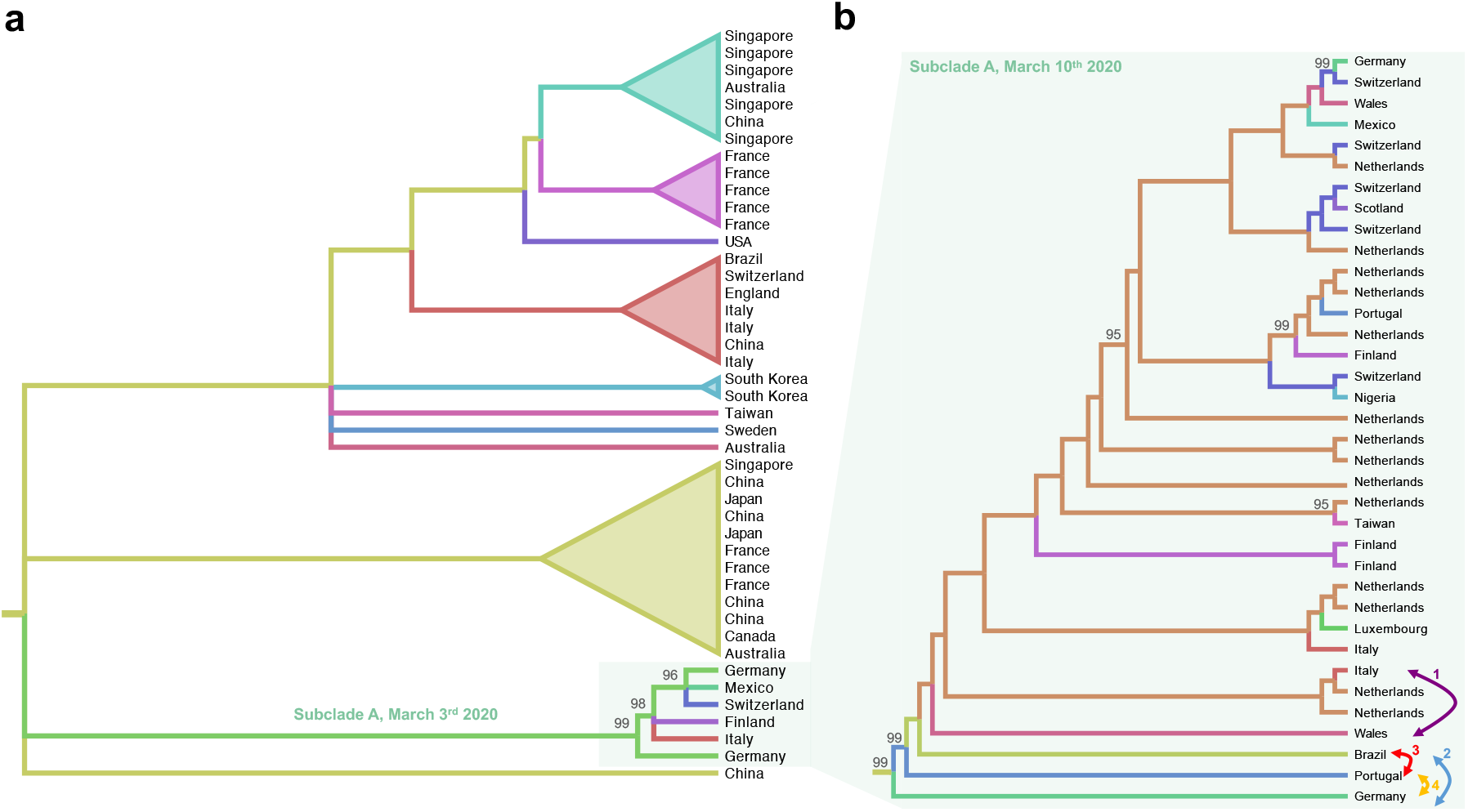
Cladograms of SARS-CoV-2 subclades. Cladograms were extracted from ML phylogenies rooted by enforcing a molecular clock. Colored branches represent country of origin of sampled sequences (tip branches) and ancestral lineages (internal branches). Numbers at nodes indicate ultrafast bootstrap (BB) support (only >90% values are shown). (a) Cladogram of a monophyletic clade within the SARS-CoV-2 ML tree inferred from sequences available on March 3^rd^ 2020 (Supplementary Figure S1). The subclade including sequences from Italy and Germany, named Subclade A, is highlighted. (b) Cladogram of subclade A of the SARS-CoV-2 ML tree including additional sequences available on March 10^th^ 2020 (Supplementary Figure S2). Each bi-directional arrow, and corresponding number, connects two tip branches that were switched to generate an alternative tree topology to be tested (see Table 1, Methods).

Based on this observation (which was available on NextStrain), a heated discussion circulated on social media about a transmission event from Germany to Italy followed by further spread from Italy the other countries. However, in a new tree inferred just one week later, when more than 135 new full genome sequences were made available on GISAID ^8^ (Figure S2), the direct link between Germany and Italy has disappeared due to the additional clustering of previously unsampled sequences from Portugal, Brazil, Wales and Netherland (Figure 2b). Most importantly, there is no sufficient evidence of a higher likelihood (Shimodaira-Hasgawa test, Table 1) for tree topologies generated arbitrarily switching branches in the tree (see arrows in Figure 2b) compared to the ML tree inferred from the real data. In other words, it is not possible with the present data to decide which branching pattern (and, therefore, phylogeographic reconstruction) is the most likely.

**Table 1.**
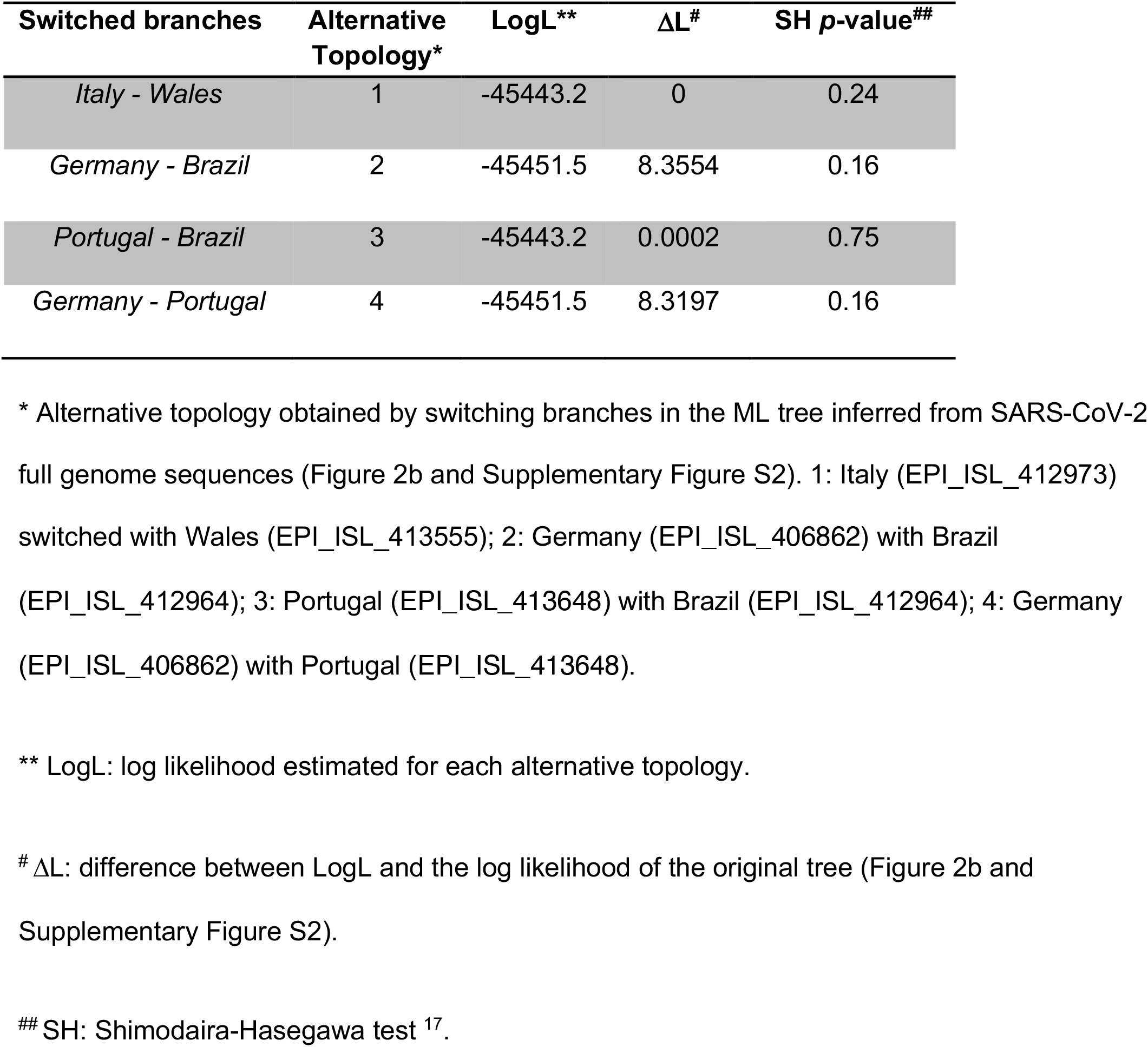
Testing alternative topologies.

## Sequencing errors and phylogenetic noise

Lack of resolution and uncertainty in the SARS-CoV-2 phylogenetic tree is to be expected, considering that relatively little genetic diversity can be accumulated during the first three months of an epidemic, even for an exponentially spreading and fast-evolving RNA virus. Plots of pairwise transitions/transversions *vs*. genetic distance ^10^ showed, indeed, low genetic diversity among SARS-CoV-2 genomes, considerable scattering, and overlap in transitions and transversions among closely related sequences (Figure 3a). The 331 full genomes available contained 477 out of 29,564 variable sites (1.6%), of which 377 were singleton and only 100 (0.34%) parsimony informative, indicating a substantial lack of phylogenetic information, and raising the possibility of a number of sequencing errors. Further assessment of phylogenetic signal in the dataset was carried out using likelihood mapping analysis ^11^, which estimates the likelihood of each of possible tree topology for any group of four sequences (quartet), randomly chosen from an alignment, and reports them inside an equilateral triangle (the likelihood map) where the corners represent distinct tree topologies and the center star-like trees. Quartets are considered “resolved” when the three likelihoods are significantly different (phylogenetic signal, most dots equally distributed in the corners, i.e. data are suitable for robust phylogeny inference), unresolved or partially resolved, when all three likelihood values or two of them are not significantly different (phylogenetic noise, most dots in sides or center areas, i.e. data may not be sufficient for robust phylogeny inference). Extensive simulation studies have shown that, for sequences to be considered robust in terms of phylogenetic signal, side/center areas of the likelihood mapping must include <40% of the unresolved quartets ^12^, way lower than the 52.9% unresolved quartets detected in the SARS-CoV-2 full genomes alignment (Figure 3b). It is important to keep in mind that such a lack of phylogenetic signal makes the overall topology of any SARS-CoV-2 tree unreliable, and even clades with high bootstrap values should be interpreted with extreme caution. Finally, the mean genetic distance of each sequence from the root of a ML tree indicated that the available genomic data lack temporal signal (no significant linear correlation between accumulated genetic distance and sampling time, Figure 3c). This is, again, expected in genomes obtained over a very short period of time (∼ three months) since the beginning of the outbreak. Bayesian analysis^13^, which infers phylogenetic and phylogeographic patterns from a posterior distribution of trees, might facilitate comparisons about different evolutionary scenarios, help in retrieving the correct topology, and estimate an accurate evolutionary rate. With more genome sequences sampled from different time points and geographic areas made available, literally, every day, Bayesian analysis may soon be a viable option. However, as long the new data do not increase phylogenetic and temporal signal, Bayesian phylogenetic and phylogeography results would remain equally questionable.

**Figure 3.**
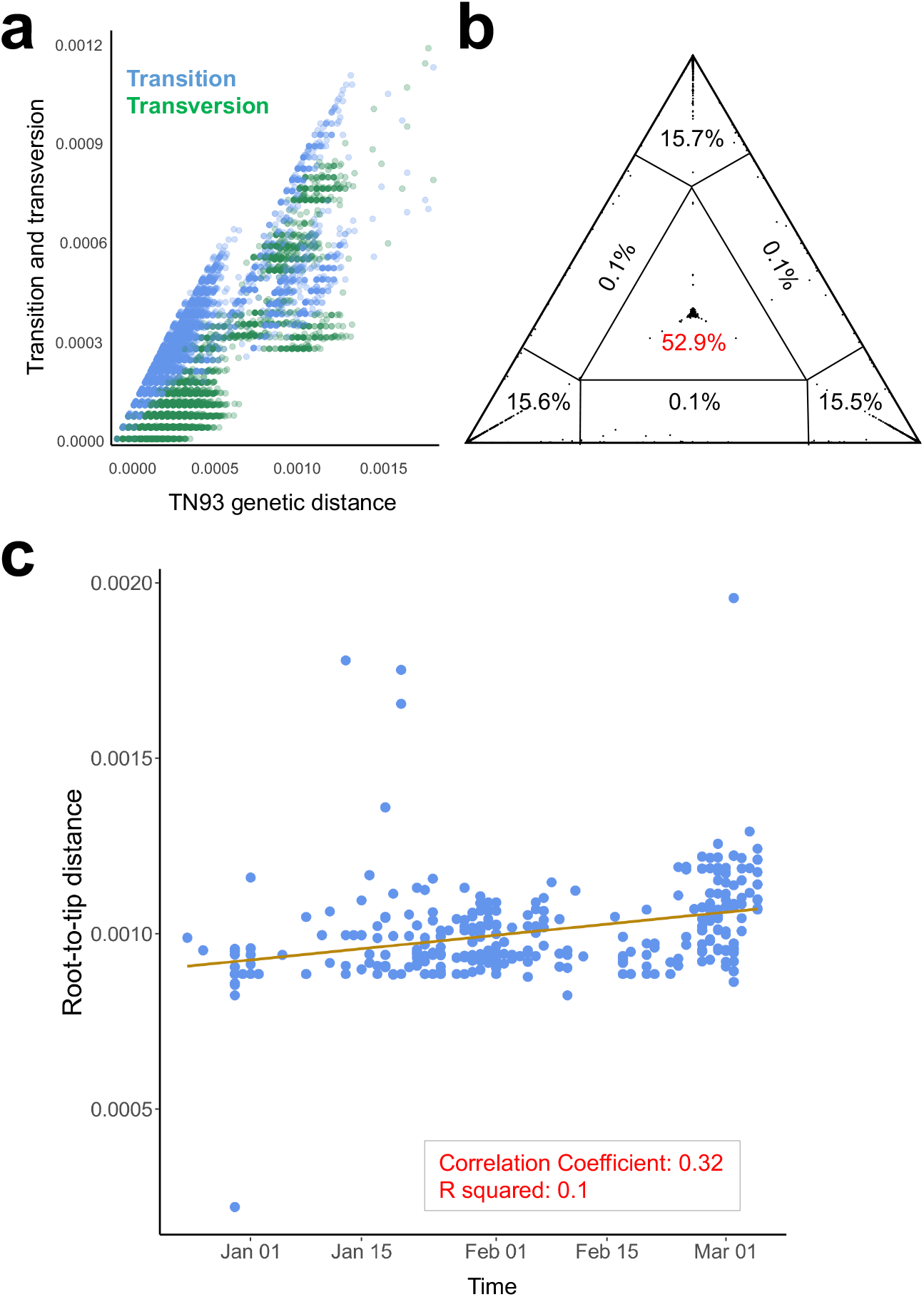
Phylogenetic and temporal signal analyses. (a) Transition (blue) and transversion (green) nucleotide substitutions (y-axis) *vs*. pairwise genetic distances (Tamura and Nei 1993 model). (b) Presence of phylogenetic signal was evaluated by likelihood mapping checking for alternative topologies (tips), unresolved quartets (center) and partly resolved quartets (edges) for the data set. (c) Linear regression of root-to-tip genetic distance in a maximum likelihood phylogeny (y-axis) *vs*. sampling time (x-axis) for each SARS-CoV-2 sequence.

## Conclusions and Future directions

Taken together, our observations suggest that any conclusion drawn, at present, about existing lineages and direction of viral spread, based on phylogenetic analysis of SARS-CoV-2 sequence data, is potentially misleading. Nowadays, published scientific data and media are very easily accessible to a worldwide audience, thus it is important more than ever to weigh the information being shared. We believe that current molecular epidemiology data are not solid enough to provide a scientifically sound analysis of SARS-CoV-2 spread. Any inference based on phylogenetic trees should be considered at the very best preliminary, and hypothesis-generating. Hence the need for 1) avoiding stigmatization based on partial information and 2) continuing the concerted scientific effort to increase the number and quality of available data. The evolutionary dynamics of SARS-CoV-2 spread will unveil an unprecedented amount of information, which is essential to make policy decisions. The whole of humanity is threatened by the current pandemic, and policy makers need to adjust their mitigation measures while the pandemic itself is developing. Some of the urgent answers required lie in the timely availability of abundant, high quality sequence data not only from countries experiencing a high number of reported cases, such as Italy and South Korea, but also from others that seem to be experiencing, at least for now, a lower number of infections.

## Methods

### Data

GISAID was accessed on March 3^rd^ and 10^th^, 2020 (final dataset information reported in Table S1). After quality control of sequences that were not full genomes or contained extensive stretches of unknown nucleotides, the first dataset (March 3^rd^ 2020) consisted of 169 genomes from 22 countries, while the second (March 10^th^, 2020) 331 from 29 countries. Confirmed cases were retrieved for the same day from interactive web-based dashboard developed by Johns Hopkins University ^14^.

### Phylogenetic signal and ML phylogeny inference

Transition/transversions vs. genetic distance plot were calculated using DAMBE6 ^10^. Evaluation of the presence of phylogenetic signal satisfying resolved phylogenetic relationships among sequences was carried out with IQ-TREE, allowing the software to search for all possible quartets using the best-fitting nucleotide substitution model ^11^. ML tree reconstruction was performed in IQ-TREE based on the best-fit model chosen according to Bayesian Information Criterion (BIC) ^15,16^. Topology was tested performing the Shimodaira-Hasegawa^17^ on “Subclade A” by comparing the inferred tree to three trees where two branches: (Tree 1) Italy (EPI_ISL_412973) switched with Wales (EPI_ISL_413555), (Tree 2) Germany (EPI_ISL_406862 with) Brazil (EPI_ISL_412964), (Tree 3) and Portugal (EPI_ISL_413648) with Brazil (EPI_ISL_412964), (Tree 4) Germany (EPI_ISL_406862) with Portugal (EPI_ISL_413648). All tests performed 10,000 re-samplings using the using the RELL approximation method ^18^. Position of these genomes within “Subclade A” is indicated in Figure 1 with an arrow.

Exploration of temporal structure, i.e. presence of molecular clock in the data, was assessed by regression of divergence -root-to-tip genetic distance-against sampling time using TempEst ^19^. In this case, absence of a linear trend indicates that the data does not contain temporal signal and that the data is not appropriate for phylogenetic inference using molecular clock models.

Ancestral state reconstruction of full datasets reported in Figures S1 and S2 were based on the ML tree obtained with IQ-TREE and performed using TreeTime ^20^. We choose to show the “European subclade” and “Subclade A” as cladograms in Figure 1 as there are no phylogenetic and temporal signals in the datasets: scaling phylogenies in time or nucleotide substitution in this case is misleading.

## Data Availability

Genomes were downloaded from GISAID
https://www.gisaid.org/, a list of the genomes used is available in the supplementary material.
Data about confirmed cases were obtained from
"Coronavirus COVID-19 Global Cases" by the Center for Systems Science and Engineering (CSSE) at Johns Hopkins University (JHU)

https://www.gisaid.org/

https://github.com/CSSEGISandData/COVID-19

## Acknowledgments

MS is supported in part by the Stephany W. Holloway University Chair in AIDS Research. IC was a member of the Italian Parliament (2013-2016). We thank all those who have contributed SARS-CoV-2 genome sequences to the GISAID database (https://www.gisaid.org/).

## Supplementary figures and tables

**Figure S1.**
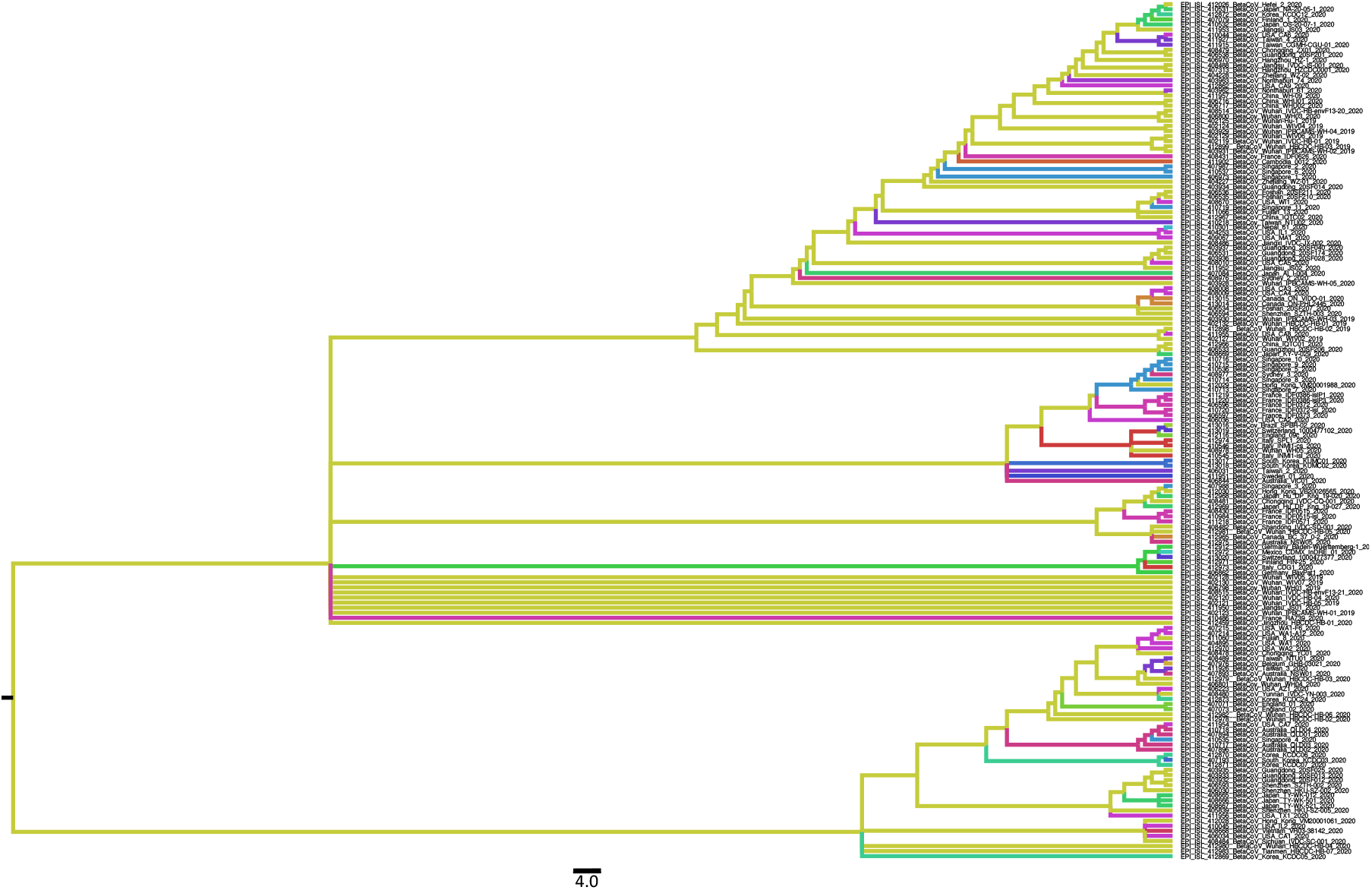
Phylogeographic reconstruction of SARS-CoV-2 across the world. Ancestral state reconstruction performed with TreeTime on 169 full genomes of SARS-CoV-2 collected on March 3^rd^ 2020. Branches are colored by countries of origin.

**Figure S2.**
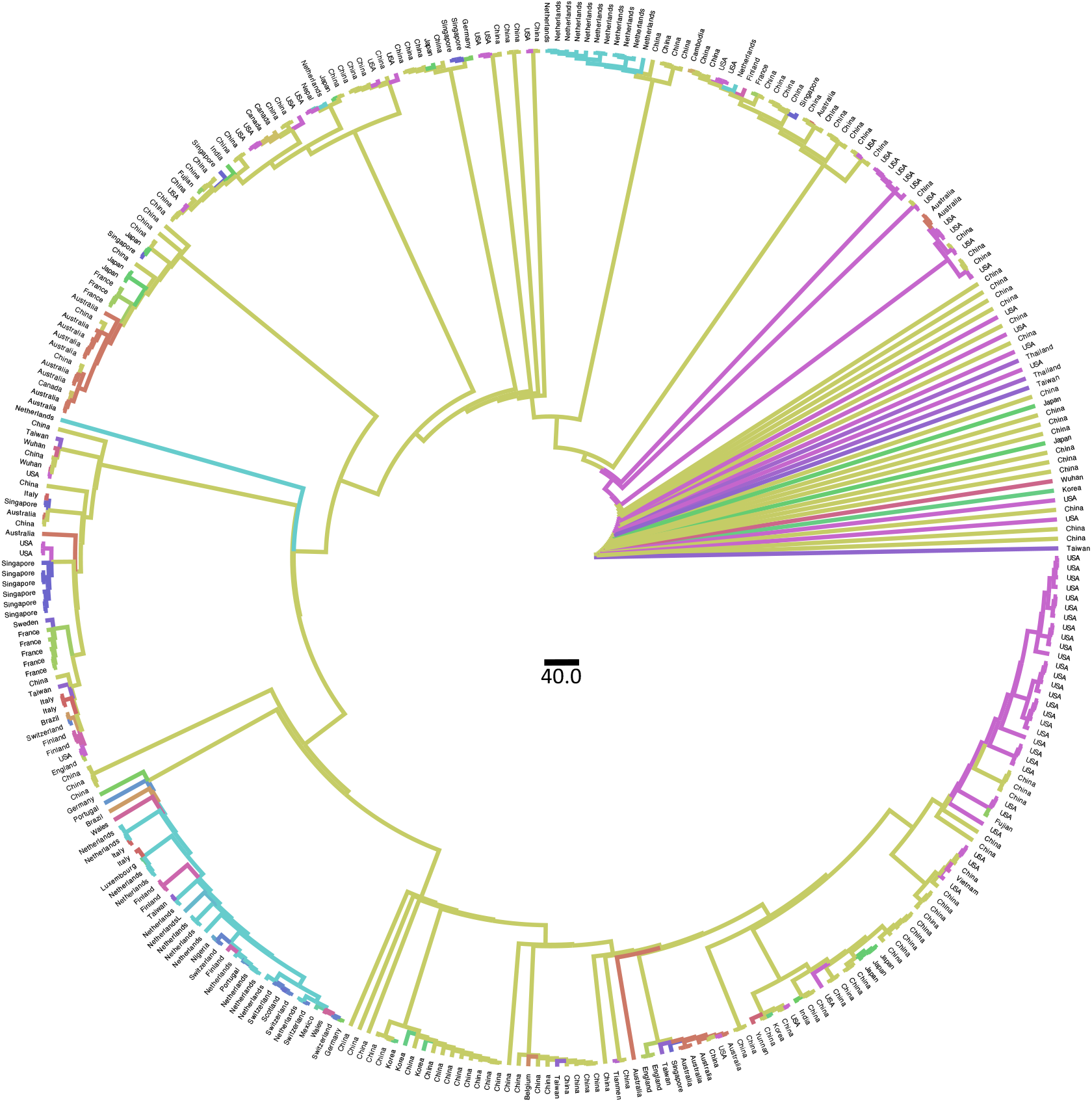
Phylogeographic reconstruction of SARS-CoV-2 across the world. Ancestral state reconstruction performed with TreeTime on 331 full genomes of SARS-CoV-2 collected on March 10^th^ 2020. Branches are colored by countries of origin.

**Table S1. Acknowledgment table with full information of genome sequences**. Downloaded from GISAID on March 10^th^ 2020 and provided as Excel file.

## References

1 WHO. World Health Organization. Coronavirus disease 2019 (COVID-19). Situation Report – 43. [online] < https://www.who.int/docs/default-source/coronaviruse/situation-reports/20200303-sitrep-43-covid19.pdf?sfvrsn=2c21c09c_2 Accessed 4 Mar. 2020]. > (2020).

2 Poon, L. L. M. & Peiris, M. Emergence of a novel human coronavirus threatening human health. Nat Med, doi:10.1038/s41591-020-0796-5 (2020).

3 Hadfield, J. et al. Nextstrain: real-time tracking of pathogen evolution. Bioinformatics 34, 4121–4123, doi:10.1093/bioinformatics/bty407 (2018).

4 Tang, X. et al. On the origin and continuing evolution of SARS-CoV-2. National Science Review, doi:10.1093/nsr/nwaa036 (2020).

5 MacLean, O. A., Orton, R., Singer, J. B. & Robertson, D. L. Response to “On the origin and continuing evolution of SARS-CoV-2”, <http://virological.org/t/response-to-on-the-origin-and-continuingevolution-of-sars-cov-2/418> (2020).

6 Kupferschmidt, K. Mutations can reveal how the coronavirus moves—but they’re easy to overinterpret, <https://www.sciencemag.org/news/2020/03/mutations-can-reveal-howcoronavirus-moves-they-re-easy-overinterpret> (2020).

7 Salemi, M., Vandamme, A.-M., & Lemey, P. The phylogenetic handbook: A practical approach to phylogenetic analysis and hypothesis testing., (Cambridge University Press, 2009).

8 Shu, Y. & McCauley, J. GISAID: Global initiative on sharing all influenza data - from vision to reality. Euro Surveill 22, doi:10.2807/1560-7917.ES.2017.22.13.30494 (2017).

9 Sito del Dipartimento della Protezione Civile - Emergenza Coronavirus: la risposta nazionale, <http://arcg.is/C1unv > (2020).

10 Xia, X. & Xie, Z. DAMBE: software package for data analysis in molecular biology and evolution. J Hered 92, 371–373 (2001).

11 Schmidt, H. A., Strimmer, K., Vingron, M. & von Haeseler, A. TREE-PUZZLE: maximum likelihood phylogenetic analysis using quartets and parallel computing. Bioinformatics 18, 502–504 (2002).

12 Strimmer, K. & von Haeseler, A. Likelihood-mapping: a simple method to visualize phylogenetic content of a sequence alignment. Proc Natl Acad Sci U S A 94, 6815–6819, doi:10.1073/pnas.94.13.6815 (1997).

13 Drummond, A. J. & Rambaut, A. BEAST: Bayesian evolutionary analysis by sampling trees. BMC Evol Biol 7, 214, doi:10.1186/1471-2148-7-214 (2007).

14 Dong, E., Du, H. & Gardner, L. An interactive web-based dashboard to track COVID-19 in real time. Lancet Infect Dis, doi:10.1016/S1473-3099(20)30120-1 (2020).

15 Nguyen, L. T., Schmidt, H. A., von Haeseler, A. & Minh, B. Q. IQ-TREE: a fast and effective stochastic algorithm for estimating maximum-likelihood phylogenies. Mol Biol Evol 32, 268–274, doi:10.1093/molbev/msu300 (2015).

16 Trifinopoulos, J., Nguyen, L. T., von Haeseler, A. & Minh, B. Q. W-IQ-TREE: a fast online phylogenetic tool for maximum likelihood analysis. Nucleic Acids Res 44, W232–235, doi:10.1093/nar/gkw256 (2016).

17 Shimodaira, H. & Hasegawa, M. Multiple comparisons of log-likelihoods with applications to phylogenetic inference. Molecular Biology and Evolution 16, 1114–1116 (1999).

18 Kishino, H., Miyata, T. & Hasegawa, M. Maximum likelihood inference of protein phylogeny and the origin of chloroplasts. Journal of Molecular Evolution volume 151–160 (1990).

19 Rambaut, A., Lam, T. T., Max Carvalho, L. & Pybus, O. G. Exploring the temporal structure of heterochronous sequences using TempEst (formerly Path-O-Gen). Virus Evol 2, vew007, doi:10.1093/ve/vew007 (2016).

20 Sagulenko, P., Puller, V. & Neher, R. A. TreeTime: Maximum-likelihood phylodynamic analysis. Virus Evol 4, vex042, doi:10.1093/ve/vex042 (2018).

